# Epidemiological behavior of the contamination curve by COVID-19 in Brazil: a time series study

**DOI:** 10.1101/2022.04.25.22274291

**Authors:** Thiffany Nayara Bento de Morais, Ketyllem Tayanne da Silva Costa, Gustavo Nepomuceno Capistrano, Fábia Barbosa de Andrade

## Abstract

The Brazil is experiencing the greatest episode of sanitary collapse ever known in the country’s history. Therefore, the relevance of this study is highlighted for the scientific advance on the epidemiological behavior of the virus in Brazil, enabling the development of analyses and discussions on the factors that influenced the high rates of contamination by SARS-CoV-2 in the country. Given the above, the study in question aims to analyze the epidemiological behavior of the contamination curve by COVID-19 by epidemiological week (EW), in the years 2020-2021, in Brazil. This is an ecological study of time series, prepared using information collected through secondary means. The country of origin of the study is Brazil, and its main theme is the number of those infected during the pandemic of COVID-19, this being the dependent variable of the study. The data been analyzed from February 23, 2020, when the first case was confirmed in Brazil, to January 1, 2022. In 2021, the country’s graph shows an exorbitant growth, reaching a peak of approximately 250 new cases per 100,000 inhabitants in the 12th EW. This data became the highest rate of the pandemic in Brazil, and did not vary significantly for the next twelve weeks. Thus, it was identified that Brazil was severely impacted by the new coronavirus, considering the high rates of confirmed cases of the virus in the country, the low adhesion of the population to preventive measures, the late start of mass vaccination in the Brazilian population, and the lack of structure in the health system, which was not properly prepared for the high demand generated by COVID-19.

## Introduction

SARS-CoV-2, the virus that causes COVID-19, was first reported in late December 2019 in Wuhan, Hubei Province, China, but soon spread to other countries. The rapid spread plus the virulence of the new coronavirus led the World Health Organization (WHO) to declare the outbreak a public health emergency of international concern and subsequently a pandemic on March 11, 2020 ^[1,2]^.

COVID-19 mainly causes respiratory symptoms, which can progress to pneumonia, and also leads to other systemic complications, such as cardiovascular, metabolic, and renal. In addition, many patients require hospitalization and, in some cases, intensive care ^[3]^.

COVID-19 has already spread throughout the world, impacting nearly 190 countries, with the highest incidence across America, Europe, and Southeast Asia. According to data from the dashboard on COVID-19 developed and managed by Johns Hopkins University (2022), as of February 22, 2022, the new coronavirus had infected 427,358,056 people and led 5,904,723 to death^[4]^.

The country that presents the highest numbers is the United States of America, occupying the first position both in those infected, with about 78.6 million, and in deaths from the disease, around 939 thousand. In the next positions are India, with about 42.8 million infected and 512 thousand deaths, and Brazil, which has 28.3 million infected, less than the number of India, but shows a higher number of deaths, which is about 645 thousand ^[4]^.

In order to try to minimize the effects of the virus, health authorities, together with governments, have instituted preventive measures, which mainly consist of the use of masks, hand hygiene, and social distancing, considering that the main means of transmission of the virus is by droplets and aerosols expelled through the respiratory tract. In cases of a positive test for the new coronavirus, it is necessary that the infected person isolates themselves, as well as the epidemiological investigation of their close contacts ^[5]^.

According to the Oswaldo Cruz Foundation (2021), Brazil is experiencing the greatest episode of sanitary collapse ever known in the country’s history^[6]^. Therefore, the relevance of this study is highlighted for the scientific advance on the epidemiological behavior of the virus in Brazil, enabling the development of analyses and discussions on the factors that influenced the high rates of contamination by SARS-CoV-2 in the country.

Given the above, the study in question aims to analyze the epidemiological behavior of the contamination curve by COVID-19 by epidemiological week, in the years 2020-2021, in Brazil.

## Materials and methods

The research in question is an ecological study of time series, prepared using information collected through secondary means. The country of origin of the study is Brazil, and its main theme is the number of those infected during the pandemic of COVID-19, this being the dependent variable of the study. The data been analyzed from February 23, 2020, when the first case was confirmed in Brazil, to January 1, 2022. The standardization of the period occurred by epidemiological weeks (WE), from the 9th EW of 2020 to the 53rd WE of 2021. The territory was analyzed considering the regions of Brazil, which show different dates of confirmation of the first case of infection.

For a better reading of the dependent variable, the morbidity rate due to SARS-COV-2 was calculated, in which the number of infected people was divided by the number of inhabitants and then multiplied by 100,000 (1). Furthermore, the independent variable for the research will be the epidemiological weeks.

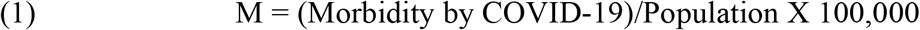

The data was collected on January 19, 2022, from a website designed by the Brazilian Ministry of Health to disseminate the COVID-19 information, and the information is updated daily and can be easily accessed via <https://covid.saude.gov.br/>.

The next step after data collection was to clean the data in Microsoft Excel® software and then perform a more rigorous analysis in JoinPoint software, version 4.9.0.0 (Surveillance Research, National Cancer Institute, USA), provided by the National Cancer Institute of the United States (http://surveillance.cancer.gov/joinpoint/) with free access and open to all public.

In the meantime, JoinPoint, through linear regression, elaborates junction points in the temporal series. Thus, two outputs are formed: the Annual Percentage Change (APC) and the Average Annual Percentage Change (AAPC), which are responsible for informing the significant variations in the analyzed rate. The Monte Carlo Permutation was the model used to demonstrate the significance in the tests, moreover, in the end the demonstration of the results will be through epidemiological curves, presenting a better view of the conduct of COVID-19 in Brazil from 2020 to 2021.

The information obtained for the construction of the study is of secondary character and was obtained through databases that have public domain, so it was not necessary to use personal data, and thus not requiring the appreciation of the Research Ethics Committee.

## Results

Table 1 and 2 demonstrate the synthesis of data achieved through linear regression due to the cases of COVID-19 in Brazil from 2020 and 2021, respectively, organized by Epidemiological Weeks (WE). Thus, it was observed in table 1 the presence of 4 JoinPoint in Brazil and in most regions, except the Midwest that had 5. In addition, Brazil presents all APC with statistical relevance, totaling 5, as well as the Southeast, but the other regions have 4. Meanwhile, in the perspective of AAPC the Midwest region presents more inconstancy in the numbers of infected and the Southeast the most constant.

**Table 1.** Morbidity rate due to COVID-19 in Brazil and regions in the period from 2020. Brazil, 2022.

| Local | Joinpoint<sup>1</sup> | Period | WPC<sup>2</sup> | Lower | Upper | AWPC<sup>3</sup> | Lower | Upper |
| --- | --- | --- | --- | --- | --- | --- | --- | --- |
| Brazil | 22, 31, 45, 50 | 9 to 22 | 48.1* | 38.9 | 57.9 | 13.3* | 10.9 | 15.8 |
|  |  | 22 to 31 | 7.8* | 5.3 | 10.3 |  |  |  |
|  |  | 31 to 45 | -6.0* | -7.0 | -4.9 |  |  |  |
|  |  | 45 to 50 | 19.3* | 11.6 | 27.5 |  |  |  |
|  |  | 50 to 53 | -9.6* | -17.8 | -0.5 |  |  |  |
| North | 19, 22, 34, 42 | 12 to 19 | 94.3* | 57.6 | 139.6 | 13.3* | 8.8 | 18.0 |
|  |  | 19 to 22 | 43.9* | 9.1 | 89.8 |  |  |  |
|  |  | 22 to 34 | -1.8* | -3.2 | -0.3 |  |  |  |
|  |  | 34 to 42 | -8.3* | -11.7 | -4.9 |  |  |  |
|  |  | 42 to 53 | 2.7 | 0.6 | 4.8 |  |  |  |
| Northeast | 22, 30, 45, 50 | 10 to 22 | 52.5* | 40.5 | 65.6 | 11.9* | 8.8 | 15.0 |
|  |  | 22 to 30 | 4.6* | 1.1 | 8.3 |  |  |  |
|  |  | 30 to 45 | -8.0* | -9.3 | -6.7 |  |  |  |
|  |  | 45 to 50 | 21.6* | 9.3 | 35.3 |  |  |  |
|  |  | 50 to 53 | -10.2 | -22.1 | 3.5 |  |  |  |
| Midwest | 25, 31, 38, 45, 50 | 10 to 25 | 52.3* | 42.6 | 62.6 | 15.8* | 12.6 | 19.1 |
|  |  | 25 to 31 | 13.0* | 7.2 | 19.0 |  |  |  |
|  |  | 31 to 38 | -1.9 | -5.3 | 1.6 |  |  |  |
|  |  | 38 to 45 | -12.4* | -16.1 | -8.4 |  |  |  |
|  |  | 45 to 50 | 10.4* | 0.7 | 20.9 |  |  |  |
|  |  | 50 to 53 | -5.6 | -17.5 | 8.0 |  |  |  |
| Southeast | 22, 31, 45, 49 | 9 to 22 | 38.9* | 30.7 | 47.6 | 11.8* | 9.3 | 14.3 |
|  |  | 22 to 31 | 9.1* | 6.3 | 12.0 |  |  |  |
|  |  | 31 to 45 | -6.2* | -7.4 | -5.0 |  |  |  |
|  |  | 45 to 49 | 21.7* | 7.7 | 37.4 |  |  |  |
|  |  | 49 to 53 | -1.1 | -7.4 | 5.6 |  |  |  |
| South | 28, 36, 41, | 11 to 28 | 33.0* | 27.4 | 38.9 | 13.9* | 11.1 | 16.7 |
|  | 51 |  |  |  |  |  |  |  |
|  |  | 28 to 36 | 7.3* | 3.2 | 11.6 |  |  |  |
|  |  | 36 to 41 | -15.7* | -23.6 | -7.1 |  |  |  |
|  |  | 41 to 51 | 17.0* | 14.2 | 19.8 |  |  |  |
|  |  | 51 to 53 | -28.9* | -43.0 | -11.4 |  |  |  |

<sup>1</sup>by epidemiological week.
<sup>2</sup>week percentage change.
<sup>3</sup>average week percentage change.
\*p-value <0.05.

**Table 2.** Morbidity rate due to COVID-19 in Brazil and regions in the period from 2021. Brazil, 2022.

| Local | Joinpoint<sup>1</sup> | Period | WPC<sup>2</sup> | Lower | Upper | AWPC<sup>3</sup> | Lower | Upper |
| --- | --- | --- | --- | --- | --- | --- | --- | --- |
| Brazil | 12,24 | 1 to 12 | 3.4* | 1.2 | 5.7 | -4.1 | -5.0 | -3.3 |
|  |  | 12 to 24 | -0.3 | -2.4 | 1.8 |  |  |  |
|  |  | 24 to 52 | -8.5* | -9.4 | -7.6 |  |  |  |
| North | 3, 7, 11, 25, 39 | 1 to 3 | 24.9 | -1.3 | 58.1 | -2.5* | -4.3 | -0.7 |
|  |  | 3 to 7 | -8.7 | -18.1 | 1.7 |  |  |  |
|  |  | 7 to 11 | 6.8 | -4.4 | 19.4 |  |  |  |
|  |  | 11 to 25 | -4.6* | 5.8 | -3.4 |  |  |  |
|  |  | 25 to 39 | -11.4* | -13.5 | -9.3 |  |  |  |
|  |  | 39 to 52 | 5.6* | 2.4 | 8.9 |  |  |  |
| Northeast | 11, 25, 32 | 1 to 11 | 6.0* | 3.5 | 8.6 | -3.3* | -4.6 | -2.0 |
|  |  | 11 to 25 | 1.2 | -0.1* | 2.6 |  |  |  |
|  |  | 25 to 32 | -22.0* | -27.0 | -16.7 |  |  |  |
|  |  | 32 to 52 | -3.6* | -5.5 | -1.6 |  |  |  |
| Midwest | 7, 10, 34 | 1 to 7 | -1.8 | -9.1 | 6.2 | -3.4 | -5.9 | -0.9 |
|  |  | 7 to 10 | 22.3 | -17.1 | 80.6 |  |  |  |
|  |  | 10 to 34 | -2.2* | -3.0 | -1.3 |  |  |  |
|  |  | 34 to 52 | -9.2* | -11.5 | -6.9 |  |  |  |
| Southeast | 25 | 1 to 25 | 1.3* | 0.1 | 2.6 | -4.3* | -5.5 | -3.1 |
|  |  | 25 to 52 | -9.0* | -10.9 | -7.1 |  |  |  |
| South | 5, 10, 14, 25 | 5 to 10 | 25.3 | 7.9 | 45.5 | -4.7 | -7.2 | -2.1 |
|  |  | 10 to 14 | -17.8 | -34.1 | 2.4 |  |  |  |
|  |  | 14 to 25 | 2.7 | -1.0 | 6.6 |  |  |  |
|  |  | 25 to 52 | -8.6* | -9.9 | -7.2 |  |  |  |

<sup>1</sup>by epidemiological week.
<sup>2</sup>week percentage change.
<sup>3</sup>average week percentage change.
\*p-value <0.05.

In relation to 2021 that has its linear regression demonstrated in table 2, Brazil presents 2 JoinPoint, the Southeast region has only one, Center-West has 3, South with 4 and North with 5 JoinPoint, demonstrating distinct behaviors. In relation to the statistical relevance of the APC in Brazil, 2 were obtained, from the 1st to 12thWE and 24th to 52ndWE. The Midwest and Southeast also had 2 APC, the Northeast demonstrated lhe greatest amount with 3 and South only with one, from the 25th to 52nd WE. When the AAPC was evaluated, the South had the most inconsistent numbers in relation to the numbers of infected by COVID-19 and the North the most constant.

Based on figure 1, it is possible to observe the behavior of the number of infected persons in Brazil from 2020 and 2021 through polynomial models. Thus, it is obtained a significant increase in the numbers infected by COVID-19 in Brazil in 2020 until approximately the 30th WE (with a rate of 160 per 100,000 people), followed by a significant decrease in cases until the 46th WE, with approximately, a rate of 80 cases per 100,000 people. However, between the 44th and 50th WE there was, again, an increase that reaches a peak with a rate of more than 170 cases, but, one more time, the behavior was accompanied by a drop until the 53rd WE that ends in 2020.

**Fig 1.**
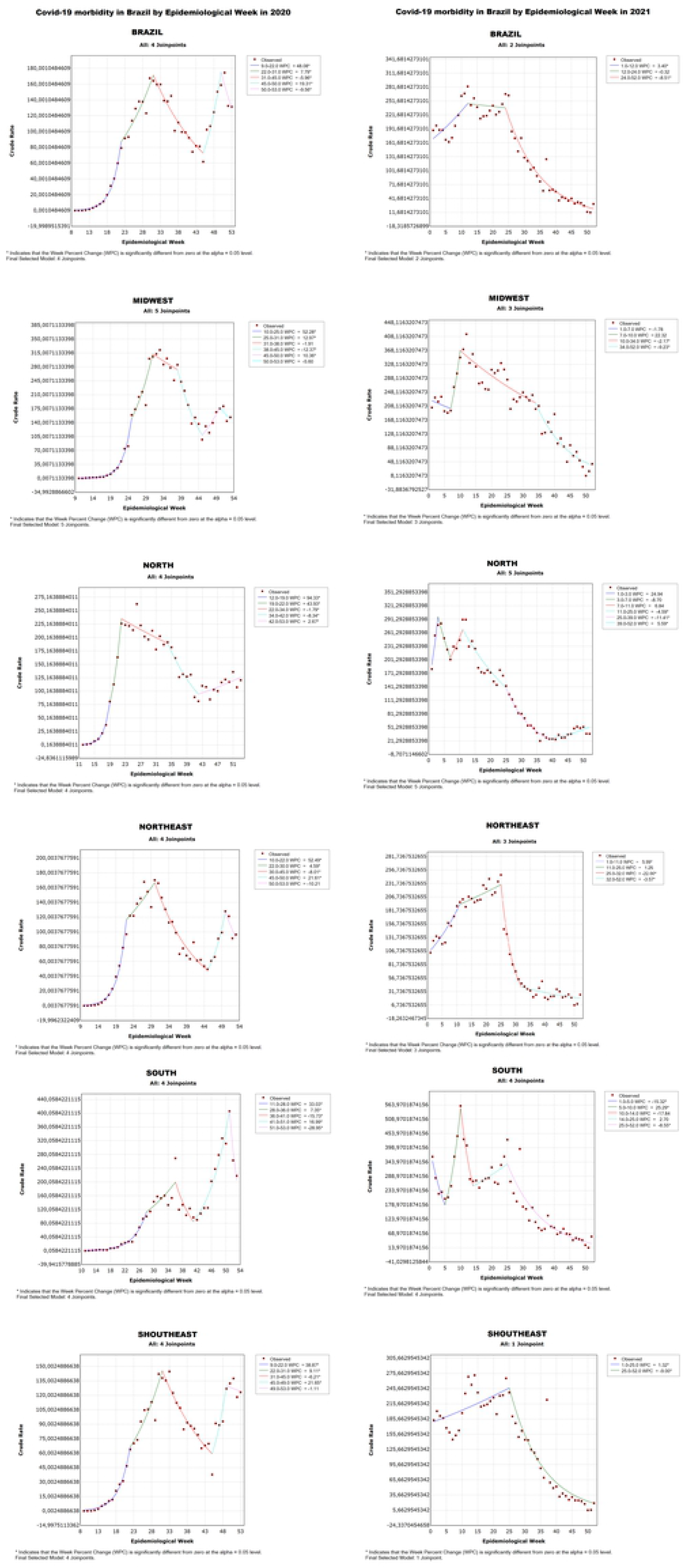
Morbidity rate due to COVID-19 in Brazil and regions in the period from 2020 to 2021. Brazil, 2022.

In this way, the Southeast, Northeast, and Midwest regions present a behavior in the ascending curve in the graph in a similar way as Brazil in 2020, with a representative increase in the rate until the 30th WE. As for the North, there was this rise in numbers until the 24th EW, with the highest rate of the region in 2020 (225 cases per 100,000 inhabitants), then there is a subtle decrease (rate of 200 in the 24th WE), but still, the North shows another decrease of those infected until the 42nd WE, with approximately 100 cases per 100,000 inhabitants and then increases the rate up to 125 cases in 51st WE.

Meanwhile, the South presents the peak in the number of cases in the 53rd WE, reaching the highest rate with more than 400 cases per 100,000 inhabitants. Still, the behavior of the drop between the 32nd and 46th WE in Brazil was similar to that of the Northeast, Southeast, and Midwest. The similar pattern occurred again, as all had at the end of 2020 an increase in the number of cases.

In addition, still in Figure 1, from the perspective of the year 2021, it is possible to see that the first 12 WE of the year shows an increase in the graph, reaching approximately the rate of 251, and this value stabilizes in the following weeks, more specifically, until the 23rd WE. However, this pattern changes with a gradual and representative drop in the number of infected that continues until the end of the year, which has the lowest rate with 12 cases per 100,000 inhabitants.

The regions that have the most similar behavior are the Northeast and Southeast, in a way that the cases in the beginning of 2021 grow until the 25th WE with a rate of 231 and 245, respectively, and then there is a accentuated decrease with a rate of approximately 7 in both regions in the 52nd WE. The Midwest starts the year with a different behavior, as there is a decrease in the numbers infected between the 0th and 7th WE with a rate of 208, then there is a considerable increase, demonstrated by a steep upward curve over the course of 3 weeks, reaching the region’s 2021 peak with a rate of 368. However, afterwards, the numbers gradually drop, once again, until the 52nd WE of 2021.

Another region that shows an analogous pattern at the beginning of 2021 is the South, as the cases begin this period with a decrease until the 5th WE with 178 cases per 100,000 inhabitants, then there is an accentuated progression reaching the peak with a rate of 563 in the 10th WE. However, the numbers drop sharply until the 13th WE with a rate of just over 233 infected. This is followed by another increase, although in a more subtle way (rate of 343 in the 25th WE), but soon there is a gradual decline that lasts until the end of the year in question.

In contrast, the North, as in the graph of Brazil and the South region, begins the year 2021 with an increase in the rate until approximately the 3rd WE, demonstrating its peak with a rate of 291 cases. It is followed by a drop (rate of 201 in the 7th WE) and a further increase (rate of 261 in the 12th WE). Then the behavior is analogous to the other regions, with a considerable drop in numbers until the finalization of the WEs of the year 2021. COVID-19 started to be confirmed in Brazil in the 8th WE of 2020, more specifically in the southeast region, while other regions tested positive for the virus in the following weeks, with the northern region presenting more delayed confirmed cases (11th WE).

With this, in the graph of Brazil, presented in figure 1, a gradual growth is observed until the 15th WE, with less than one case per 100,000 inhabitants. Then, there is a period of rapid increase in the number of cases until the 30th EW, which reached more than 160 cases per 100,000 inhabitants. Afterwards, the graph shows a drastic reduction in the rate, with confirmation of approximately 70 new cases. Finally, the year ends with a rise that peaks in morbidity at more than 170 proportional cases, followed by a drop of about 20 cases in the 2020 rate.

In 2021, the country’s graph shows an exorbitant growth, reaching a peak of approximately 250 new cases per 100,000 inhabitants in the 12th WE. Thus, this data became the highest rate of the pandemic in Brazil, and did not vary significantly for the next twelve weeks. Then there is a gradual decline in the rate of confirmed cases, which reaches about 20 infected people per 100,000 inhabitants by the end of the year.

In this sense, the quick dissemination of the virus caused a collapse in the world health system, especially in underdeveloped countries like Brazil. Therefore, during the pandemic, especially during the morbidity and mortality peaks of COVID-19, the lack of supplies, beds, equipment, and skilled labor became a common problem.

## Discussion

In the year 2020, health professionals and researchers in the field faced a challenging scenario due to the high rate of infection by COVID-19, considering that this was a recent disease and with incipient studies on transmissibility pattern, infectivity, lethality and mortality, without specific and effective vaccines and drugs to treat the virus ^[7]^.

Knowing that the virus is spread mainly by droplets and aerosols expelled by the respiratory tract, measures to prevent and contain the virus were adopted, such as social distancing, with the cancellation of events and closure of public and private institutions, allowing only the operation of essential activities, such as hospitals, pharmacies and supermarkets, in addition to the recommended quarantine of the infected person and their close contacts, and hand washing ^[8]^. The use of masks was not recommended at the time, as there were not enough studies proving their effectiveness in preventing the coronavirus ^[9]^.

Even the activities in the health area underwent changes, with partial and restricted functioning. The services provided were mainly focused on coronavirus cases, which led to the reallocation of professionals from other sectors to the COVID-19 units, in order to alleviate the high demand generated by the crisis. ^[10]^. With this, many patients with chronic diseases and who were on continuous treatment were penalized due to the reduction in specialized care.

In the pandemic context, there were abrupt changes in social dynamics, with the rapid advance of the virus, causing high morbidity and mortality rates. Moreover, due to the scarcity of scientific evidence and, concomitantly, an excess of theories and information, often contradictory, caused confusion and uncertainty in the population ^[11]^.

All the factors mentioned contributed to the increasing infection of the virus in Brazil, which can be observed in figure 1, that shows an ascending curve until EW 30, reaching the peak of the period of more than 160 cases per 100,000 inhabitants. The regions of the country presented an epidemiological behavior similar to the pattern in Brazil, with the exception of the Northern region, which grew up to 225 cases per 100,000 inhabitants in WE 22, and the Southern region, which grew until WE 36, reaching the peak of the period with a rate of around 200.

Following this period of increased infection, the country experienced a reduction of confirmed cases of COVID-19 from WE 30 to 44, which is possibly related to the consolidation of more scientifically based studies on effective methods of prevention, as well as the structuring and greater investments in health, such as the construction of field hospitals, increasing the amount of free diagnostic tests for the population and the incentive to adopt preventive measures ^[12]^.

Besides public health issues, the economy has also been greatly affected by the pandemic, with increased economic vulnerability, especially in middle-income countries ^[13]^.

Another important factor in curbing the spread of the virus was the economic incentive of R$ 600.00, established by the Brazilian government through Decree No. 10.316, dated April 7, 2020, with the objective of supporting families in situations of social vulnerability, who survived on informal jobs that caused a greater exposure to the virus ^[14]^.

Even after the reduction of the epidemic curve, the country presented a new wave of infections, which led to the peak of contamination in the year 2020, in WE 49, with a rate of approximately 180 cases per 100,000 inhabitants, as shown in figure 1.

This behavior may have been influenced by the election campaign period, which was allowed by Resolution no. 23,627, of August 13, 2020, starting in September and ending on voting day, which occurred on November 15, corresponding to WE 47 ^[15]^. In the mentioned period, there were several outbreaks of agglomeration throughout the Brazilian territory, often without the adherence to preventive measures, such as the use of masks and frequent use of alcohol gel, which may have been reflected in the increase of cases.

Due to all these factors, the World Health Organization (WHO) declared South America as the epicenter of COVID-19, and Brazil became one of the most affected countries by the virus, assuming the second position in the morbidity ranking in the year 2020, behind only the United States (USA) ^[16,17]^. At the end of the same year, the pattern in some regions was the reduction of the infected rate, with the exception of the North region that continued with a growth pattern, which is maintained in the year 2021, as well as the other regions.

With the increase in morbidity, the population began to be more committed to social isolation. This adherence remained low throughout the pandemic in Brazil, reaching 52.3% in December 2020, the highest percentage since May, and a decrease in the number of confirmed cases could be observed in the last epidemiological weeks of the year. However, this was not a durable reality, as the isolation rate gradually decreased in the following year, reaching 31.1% in February/2021, reflecting the new increase in the morbidity rate by COVID-19 ^[18]^.

In the same period, the epidemiological curve in Brazil, in 2021, presents a jump in the first 12 WE, reaching the rate of approximately 251 cases per 100,000 inhabitants. Even the South, which started the year with a low perspective of infection by COVID-19, presented the peak of the pandemic in WE 10, with a rate of about 563 cases per 100,000 inhabitants. This behavior may have been influenced by Carnival, an event of Brazilian popular culture, which consists of crowded events and, even though it was forbidden due to the pandemic, there were many clandestine agglomerations in WE 7.

At that time, there were already COVID-19 vaccines on the market, with scientific proof of efficacy. However, due to high demand and issues related to commercial transactions between the countries and the vaccine manufacturing companies, Brazil started vaccination late in comparison to other countries from Europe and North America ^[19]^.

Between WE 12 and 23, the curve was stable at the peak and then showed a drop from WE 23, which corresponds to the month of June, until the end of the year. According to Our World in Data COVID vaccination data, Brazil began vaccination against COVID-19 in the second half of January 2021, as did India. However, the US has already started vaccination earlier, in December 2020. Still, what can be observed is that the speed and vaccination coverage in the US was higher, reaching the peak of daily vaccination in April/2021 with 1.06% of the total population, while Brazil only reached the highest number of vaccines applied in 24 hours in August/2021 with 0.91%, but was already showing high rates since June/2021 ^[20]^.

As shown in Figure 1, Brazil, as well as all of its regions showed a pattern of gradual decline of WE 23 until the end of 2021. In the last WE, the country shows a rate of confirmed cases of approximately 11 cases per 100,000 inhabitants and, in the same period, the vaccination percentage was 75.6% with the first dose and 67.2% with the complete vaccination scheme. These data corroborate the effectiveness of the vaccine ^[21]^.

## Conclusion

This study performed an analysis of the epidemiological behavior of the contamination curve by the new coronavirus by epidemiological week, in the years 2020-2021, in Brazil. Thus, it was identified that Brazil was severely impacted by the new coronavirus, considering the high rates of confirmed cases of the virus in the country, the low adhesion of the population to preventive measures, the late start of mass vaccination in the Brazilian population, and the lack of structure in the health system, which was not properly prepared for the high demand generated by COVID-19.

The limitations of the study include the difficulty in accessing morbidity data in the public information systems in Brazil, as well as the scarcity of variables in the public domain and easy access for a better analysis of the epidemiological profile of confirmed cases of COVID-19.

## Data Availability

All relevant data are within the manuscript and its Supporting Information files.

## Financing

This study will be partially funded by the Coordination for the Improvement of Higher Education Personnel - Brazil (CAPES) - Financial Code 001. Funders will have no role in the study design, data collection and analysis, publication decision or preparation of the manuscript.

